# *“Whenever I tell her to wear slippers, she turns a deaf ear. She never listens”* : a qualitative descriptive research on the barriers to basic lymphedema management and quality of life in lymphatic filariasis patients in a rural block of eastern India

**DOI:** 10.1101/2023.03.10.23287017

**Authors:** Pragya Kumar, Shamshad Ahmad, Ditipriya Bhar, Ria Roy, Bhavna Singh

## Abstract

**Background:** Chronic lymphatic filariasis cases in Bihar, India, need management of lymphedema to live a life free of disability. For patients who have recurrent attacks of acute dermato-lymphangio-adenitis (ADLA), WHO has recommended simple home-based measures that include maintaining hygiene, skin care, and limb movement. But patients in rural areas are unable to adopt them, resulting in a vicious cycle of ADLA attacks. So there might be multiple realities from patients’ and healthcare workers’ perspectives that are unexplored. A qualitative research was deemed best suitable to identify the barriers to practising home-based lymphedema practices that are adversely affecting quality of life.

**Methods:** The qualitative descriptive study was conducted in two villages in the rural field practice area under a tertiary care hospital in Bihar. Researchers purposively selected ten participants, including patients affected by lymphedema, their caregivers, the grassroots healthcare workers, and the block health manager. In-depth interviews were conducted using a semi-structured interview guide. Data was entered into QDA Miner Lite, where researchers did attribute, in-vivo, process, descriptive, emotion, and holistic coding, followed by content analysis, where categories and themes emerged from the codes.

**Results:** Three themes emerged: the inherent nature of disease, patient related factors, and healthcare system related factors. Besides low awareness and adherence, low health-seeking behaviour and poor personal hygiene, categories like signs and symptoms, seasonal factors, hampered activities of daily living, hopelessness from not getting cured, psychosocial difficulty, lack of capacity building and receipt of incentives by healthcare workers, unavailability of lab diagnosis and management of complications at the facility, inconsistent drug supply, and no financial assistance were the identified barriers.

**Conclusions:** Accessibility to WaSH, regular training of home-based care, increasing the capacity and motivation of grassroots workers, and the generation of in-depth awareness among the patients are required to achieve the elimination of filariasis, with MMDP as a key component of that strategy for endemic districts across the whole country.

## Background

Lymphatic filariasis (LF), a neglected tropical disease, is endemic in 257 districts of India, putting 650 million population at risk. They include all 38 districts of the state of Bihar [1]. Lymphedema is the commonest morbidity among the chronic cases. For achieving elimination of LF, two-pronged strategies are used: annual mass drug administration (MDA) with diethylcarbamazine and albendazole, and morbidity management and disability prevention of lymphedema (MMDP) [1, 2].

Under MDA, there are biannual rounds of door-to-door distribution of tablet albendazole (which can be combined with diethylcarbamazine and/or ivermectin) followed with supervised consumption of the tablets by the healthy and afflicted alike. The grassroots healthcare workers supervise the distribution of the medicines in their respective areas. Under MMDP, an essential package of care is provided to only the patients with filarial lymphedema, either by the healthcare workers or directly at the block health centre. Washing the affected limb daily with soap and clean water at room temperature, followed by drying with a clean cotton cloth is the recommended protocol. Interdigital lesions are usually treated with anti-fungal creams; proper footwear is to be used to avoid ‘entry’ lesions, while antiseptic or antibiotic creams are used for small wounds. For management of hydrocele, an important surgical complication, hydrocelectomy is the treatment of choice [3]. There is a huge burden of chronic cases in Bihar which needs management of lymphedema for making life free of disability possible. Yet these patients are not effectively being followed up under the present elimination program of LF: only 7155 MMDP kits had been distributed among the patients, and 129 hydrocelectomies done in the block health centres in Bihar in 2021 [4].

For the patients who have recurrent attacks of acute dermato-lymphangio-adenitis (ADLA) in their limbs, WHO has also recommended simple home-based measures that include maintaining hygiene, skin care and limb movement. Patients can do compressive bandaging, elevation of leg while resting, and careful washing with soap and water. But patients in our rural practice area are unable to adopt them, resulting in vicious cycle of ADLA attacks. The water, sanitation and hygiene (WaSH) component has thus become essential in the elimination of LF [5]. Despite this common knowledge, low health-seeking behaviour, poorer socio-economic conditions, and lesser adoption of personal protection are some of the known reasons behind decreased lymphedema management practices. Moreover, there might be multiple realities from the patients’ and healthcare workers’ perspectives, which have yet to be explored. Based on this background, a qualitative research was deemed best suitable to identify these barriers in practicing home-based lymphedema practices in the rural area of Bihar, which are adversely affecting the quality of life in LF patients. Finding out and building a comprehensive picture of the barriers, and understanding how these barriers affect the quality of life, would help both the patients and the healthcare system in addressing ADLA in rural areas of Bihar.

## Methods

### Study setting and design

LF is one of the six diseases covered under the National Vector Borne Disease Control Programme (NVBDCP), which is an umbrella program of the Government of India dealing with vector-borne diseases namely Malaria, LF, Dengue, Kala-azar, Chikungunya and Japanese Encephalitis. All 38 districts of Bihar are highly endemic for LF, and has the highest caseload in India. In 2021, as many as 89970 lymphedema and 19566 hydrocele cases were reported by NVBDCP in Bihar [4]. This qualitative descriptive study was conducted in two villages, Chakiyapar and Maharajganj, in the rural field practice area (Naubatpur block) under a tertiary care hospital in the state of Bihar. The researchers selected these villages under the rural field practice area because they have highly reported caseloads.

### Research members

The research was led by Pragya Kumar (PK) and Shamshad Ahmad (SA), who are senior faculty at the Department of Community and Family Medicine at a tertiary care institution of national importance in India. Both researchers were well trained and experienced for carrying out qualitative research methods. The other members of the team, Ditipriya Bhar (DB), Ria Roy (RR) and Bhavna Singh (BS), were trained in interviewing techniques, transcription, translation and coding procedures in qualitative research. All the members were fluent in the regional language, and were involved in conducting in-depth interviews with the study participants.

### Study tool

A semi-structured in-depth interview guide was developed by the researchers PK and SA for each stakeholder (Additional file 1: Text S1). The interview guide was formed initially with a deductive framework, but with room to include any new emergent theme during the process of data collection or data analysis. The questions were asked in order for the first patient and caregiver, but later modified according to severity of the health condition or any new theme detected.

### Study population

The patients of LF affected with lymphedema, their caregivers, and the healthcare workers working in the rural field practice area were considered the study population. The caregivers are unpaid members of the patients’ family or social network, who help them with the activities of daily living, and live within an hour reach from their homes. One cadre of healthcare workers considered here are grassroots healthcare workers, namely accredited social health activists (ASHA) in these villages, with one worker looking after the basic health assistance of 1000 rural population. They are the first point of contact for healthcare delivery system in the area. The other cadre is the block health manager who looks after the overall service delivery for LF to all the villages covered under the block primary health center (BPHC) of Naubatpur, including Chakiyapar and Maharajganj. All the MMDP kits and MDA drugs are supervised by the block health manager. For our study, the inclusion criteria were: (i) patients more than 18 years old, living with lymphedema for more than one year, (ii) caregivers of filariasis patients with lymphedema who were present at the patients’ residence during the study, and (iii) the grassroots healthcare workers and the block health manager of the rural field practice area.

### Study technique, data collection and analysis

Following the informed consent, PK, SA, DB and RR conducted in-depth interviews, in teams of two, using the interview guide. Each interview lasted around 8-10 min and their responses were further probed. The teams organically explored new areas of inquiry in the interviews as they emerged from the answers. The audiotape of each interview was played several times, paying close attention to remove any unnecessary phrase. The audiotapes were first transcribed with exact verbatim by DB, RR and BS, by listening to a clip every 10 sec before transcribing it. Each audiotape took around 40 min to transcribe in local language (Hindi), and then 1 h to translate to English on the same day after the interview. This helped in capturing any hesitation or emotion behind the words spoken by the participants. The names of interviewees (and of other people mentioned during their interviews) were removed from the transcripts. The researchers DB, RR and BS re-read the transcripts several times, and important aspects, which were immediately striking, were marked.

Data was collected from ten participants: four patients suffering from lymphedema, three caregivers of LF patients, two grassroots healthcare workers, and the block health manager of the rural area. The transcripts were entered into QDA Miner Lite software, where two of the researchers independently performed mainly attribute, in-vivo, process, descriptive, emotion and holistic coding. If any coding process hit a block, DB, RR and BS listened to the audiotapes again to understand the intended meanings of the participants. The data collection and data analysis happened concurrently, to revise the interview guide and draw upon the emergent areas of inquiry. The codes were shared between all the researchers, and disagreements were resolved. PK, SA, DB and RR continued to purposively sample the patients, caregivers and healthcare workers until saturation of the codes was reached by agreement among all the researchers. Then DB, RR and BS did a content analysis, where categories and themes emerged from the codes. In this way, they identified the categories pertaining to lymphedema management, and uncovered the relationships between categories by iterative reading. Then they harmonized the themes and reconciled any matter of disagreement. Finally, a thematic codebook was made as the final coding template, that was re-applied to the data to check consistency.

## Results

The researchers tried to elucidate the factors responsible for the decrease in quality of life of the LF patients in the study through in-depth interviews. Three themes emerged in this study that provided detailed descriptions of the findings: “inherent nature of disease”, “patient-related factors”, and “healthcare system-related factors” (Fig. 1). The factors that were already assumed in the epistemology, such as low awareness and adherence, low health-seeking behaviour, and not maintaining personal hygiene, either due to poverty or non-use of WaSH facilities, are shown differently than the newer categories that emerged under patient-related factors.

**Fig. 1.**
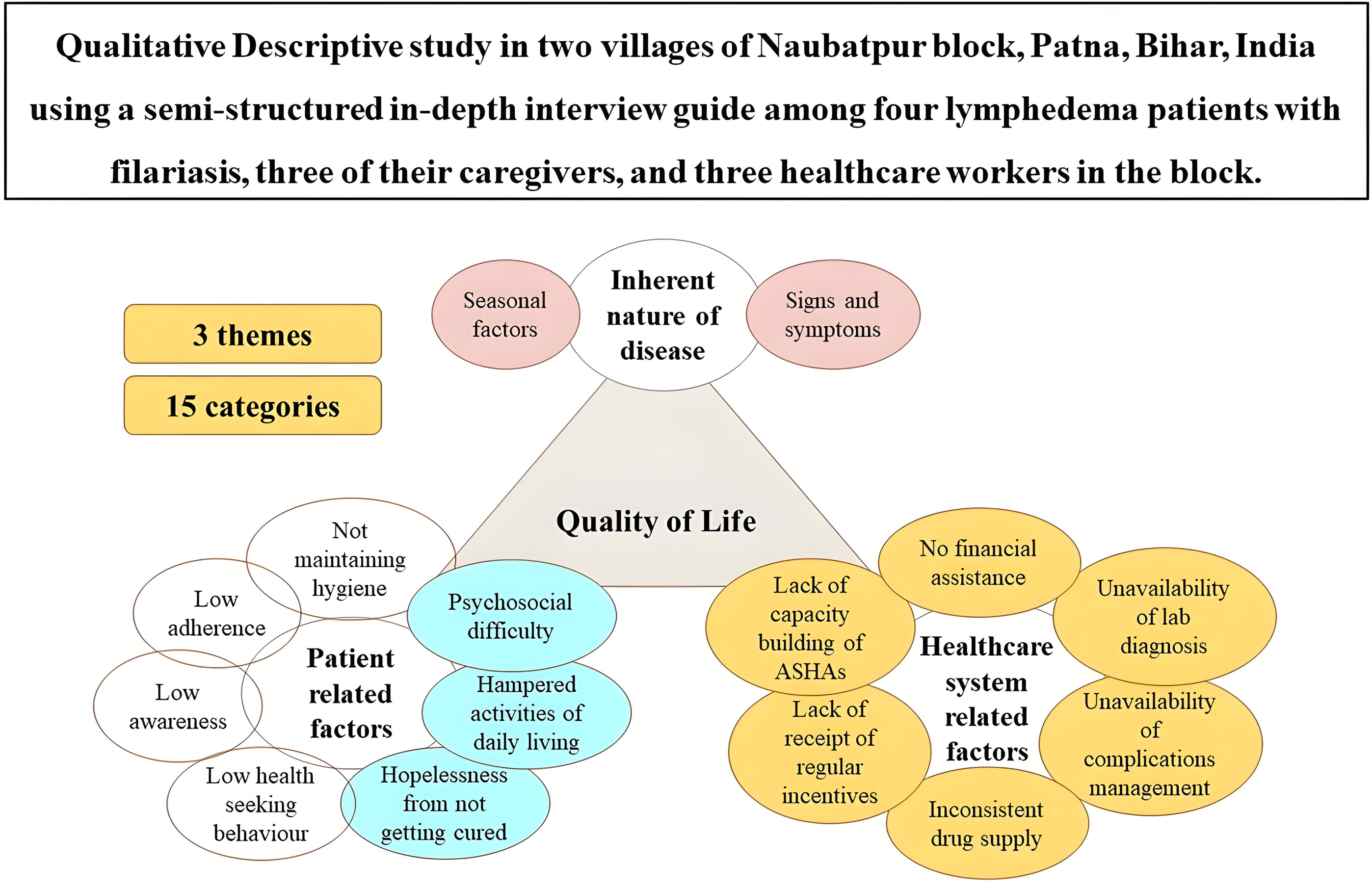
Categories and themes from the qualitative descriptive analysis.

The inherent nature of disease played a major part in the difficulty faced by participants, as evident by the frequency of its appearance in the transcripts (Table 1). The codes under signs and symptoms occurred 55 times (35.3%), while seasonal factors also accounted for 14 (9%) codes among participants. Following the category of not maintaining personal hygiene (18, 11.5%), low awareness (14, 9%), and low adherence had the greatest number of mentions among patients (12, 7.7%). The healthcare worker cadres complained about the lack of capacity building (3, 1.9%) and twice as much about the lack of receipt of regular incentives (4, 2.6%). However, most participants complained about the unavailability of laboratory diagnosis and no financial assistance (6.8%). Inconsistent drug supply had been brought to notice at least four times (2.6%) by both patients and healthcare workers.

**Table 1.**
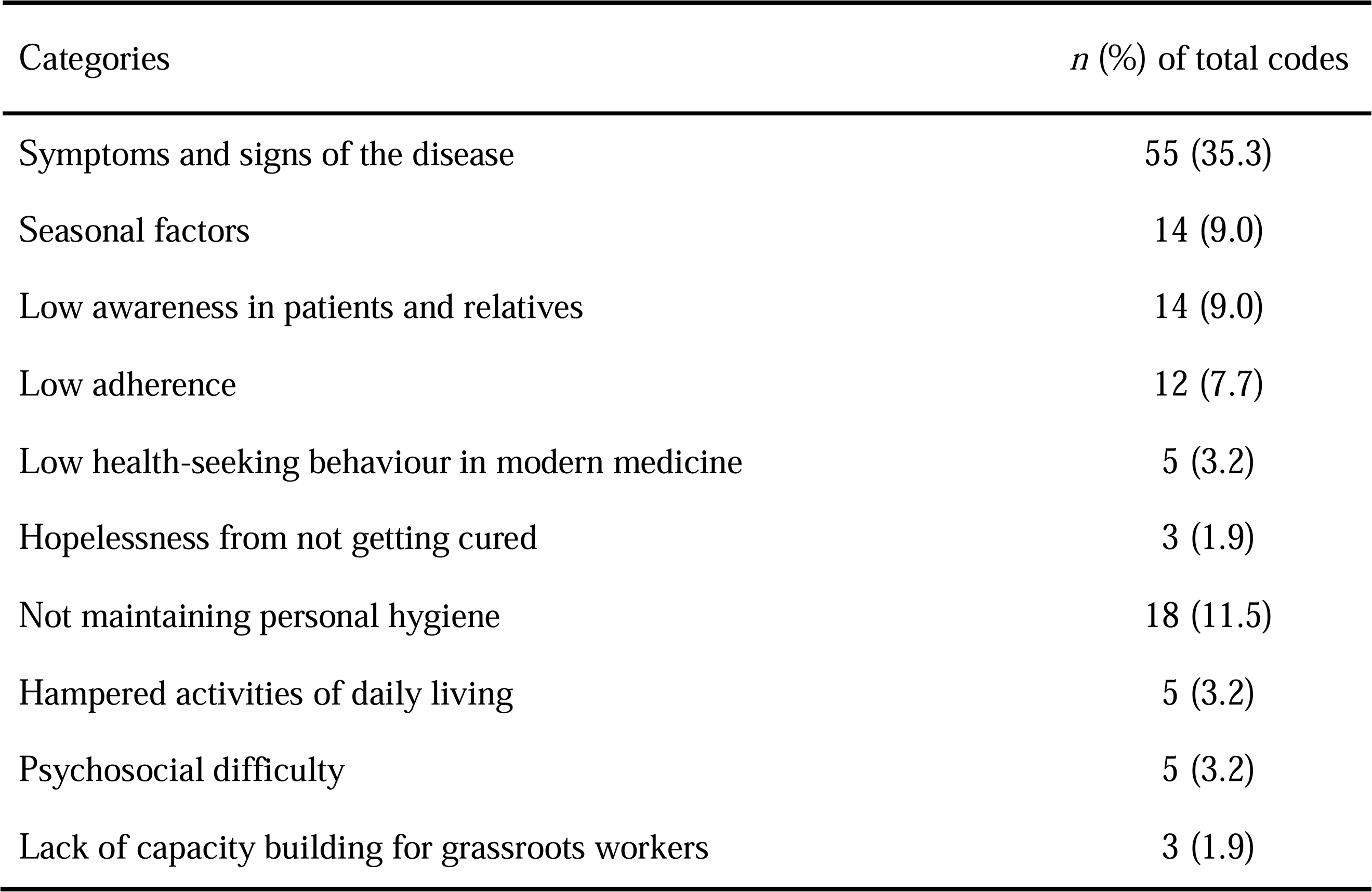
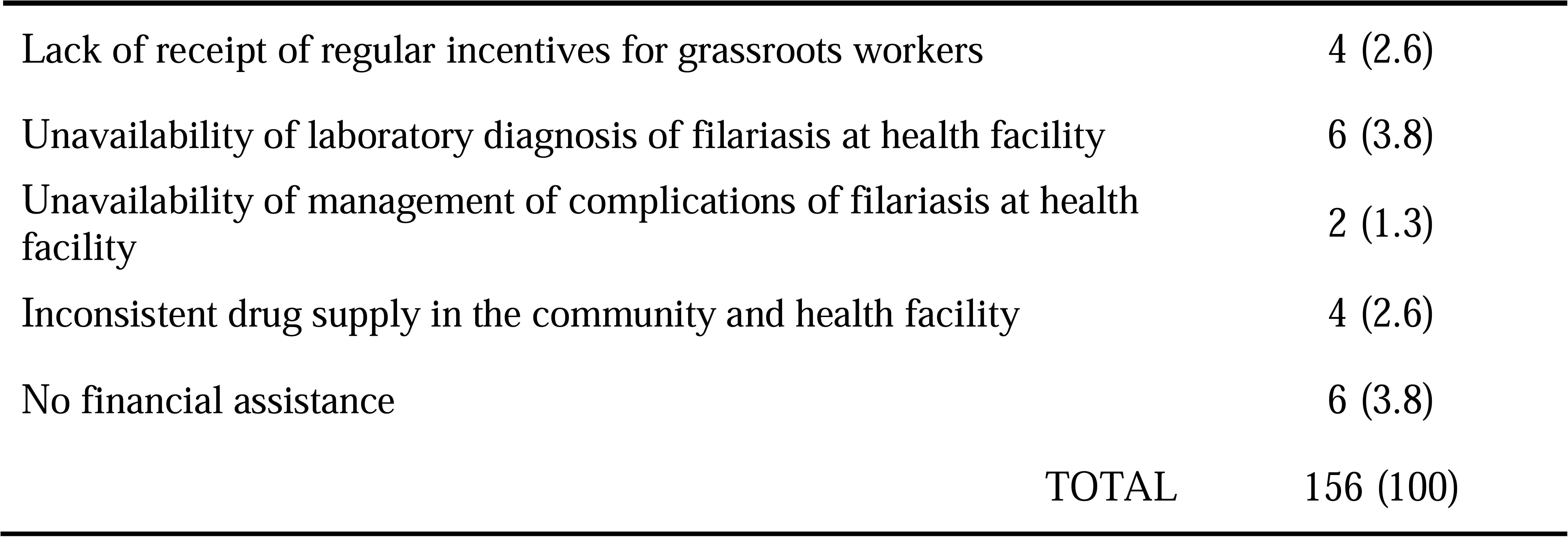
Content analysis of the categories in the study.

### Inherent nature of disease (theme 1)

#### Symptoms and signs of the disease

Patients had experienced fever, pain, and discomfort in their whole body; pain, burning sensations, and swelling in the lower limbs, including the ankles; red eyes; vomiting; weakness; and complications such as hydrocele. Thus, these multiple symptoms made the patients unable to maintain their health and increased their hardships in life, as quoted by two participants below:

> *“In a month, fever comes four times. Only on sweating it becomes alright.” **(patient, homemaker in her 40s)***

> *“Once fever came. It was so severe that my legs were swelled up, it was even more severe compared to current swelling. And blisters appeared … it was like blisters that appear after burn…. There was severe burning sensation, that I couldn’t sleep due to this.” **(patient, Anganwadi worker for 15 years)***

All these symptoms not only had a great effect on the quality of life but also portrayed symptoms that caused severe suffering due to ADLA. Their persistence over multiple years had added to the difficulty faced during the disease. Sometimes the disease was resistant to the usual treatment, and this made the condition chronic over the years, as quoted by the patient:

> *“In 2013, I went to see the doctor (at) hospital… they prescribed an injection from there. I do not remember the name. They gave 12 injections daily at morning and evening. At that time some swelling diminished, but after that gradually swelling appeared (again).” **(patient, Anganwadi worker for 15 years)***

#### Seasonal factors

Bihar has a very high temperature of around 40 degrees Celsius during the summer season, a heavy monsoon during the months of June to September, and a harsh winter ranging from November to February. A wide variation in the occurrence of ADLA has been noted, with difficulty mainly occurring during the summer.

> *“In winter discomfort used to be less, but in summer the discomfort increases. Swelling also increases in summer. The burning sensation in my legs become more severe and sometimes my whole body becomes swollen… in one leg, swelling persists throughout the year but in another leg, swelling occurs only in the summer. During winter season, this leg (showing her leg that swells in summer and currently swollen) becomes normal, but in summer it becomes like this (by showing her other swollen leg).” **(patient, Anganwadi worker for 15 years)***

There was reporting of a high fever and a flare-up of swelling in the lower limbs in two patients during the winter. The cold weather is unfavorable for the development of microfilariae but favorable for the breeding conditions of the vector mosquito, and vice versa. This results in more injection of microfilariae into the bloodstream and accumulation of the pathogen in the lymphatic system, which may explain the discomfort felt by these patients in our study during the winter.

> *“(Swelling of leg) creates more problem for him in winter, and less in summer… He doesn’t have any problem doing anything in summer. But if he overexerts himself, then everything swells up.” **(caregiver of vegetable seller in her 30s)***

### Patient related factors (theme 2)

#### Low awareness in patients and relatives

The participants had some basic knowledge about the nature of the disease; however, around half of the respondents did not have awareness about prevention and treatment available in the community, and the health facilities were lacking. One respondent reasoned that she was not formally educated, even though she was an Anganwadi worker.

> *“Not much information about filaria… Nowadays, social media is there. Like, whenever pain is there, I open YouTube and see, what could be done.” **(caregiver, a hospital attendant)***

The caregiver was also unaware about how to maintain hygiene of the affected limb of the patient:

> *“Whenever I tell her to wear slippers, she turns a deaf ear. She never listens… (I know) nothing regarding cleaning her leg.”*

Regarding the MDA programme, their low trust in government services had led them to find other options, one of which was buying over-the-counter medicines and self-medicating without a doctor’s prescription. They had been slowly discouraged from taking the MDA supply because of false information about their effectiveness, which has clearly deterred the implementation of the programme.

> *“I have not been to any doctor. (I have taken) medicines, like, these small tablets, something like that.” **(patient, farmer in his 40s)***

> *“No, I didn’t take it. Earlier where I used to live, I had taken the medicines. I remember. I ate once or twice the drug for Filaria. You know, ASHA and ANMs; they only gave the drugs.” **(caregiver, a hospital attendant)***

> *“Some people go to medical shops and show their drug there and that shop owner says that the drug is of no use and he gives another drug telling his drug would work better.” **(health care worker)***

#### Low adherence

The low adherence was in relation to preventive chemotherapy (MDA). There was a lack of trust in the government’s supply of free MDA drugs. Some revealed straightaway that they did not consume the medicine given to them biannually by healthcare workers. This problem has been shown from a patient’s, a relative’s, and a health care worker’s perspective:

> *“I was taking another medication, how could I take another? When they gave medicine (MDA), I thought I will take, but the same night I got fever and chills.” **(patient, homemaker in her 70s)***

> *“He doesn’t listen (to anyone). He says it (MDA) doesn’t work. Taking only those medicines (from private) work. If it’s for deworming, then he takes (from Government supply).” **(caregiver of vegetable seller in her 30s)***

> *“When I meet those people, I ask them whether they took the drug or not… few reply yes while rest say no…most people don’t take it in front of me, and I don’t know what they did with the drug.” **(health care worker)***

#### Low health-seeking behaviour in modern medicine

There was an overall low health-seeking behaviour among the patients. It could have been due to self-medication, as mentioned before. It could also be rooted in some barriers to receiving adequate treatment, such as the high cost of private healthcare services. Some patients tried alternative therapy for filarial lymphedema. But they eventually faced failure in getting cured, which has further aggravated their suffering:

> *“I tried in ayurvedic, allopathy, homeopathy. Everywhere, but it wasn’t successfully treated… no medicines are available. I don’t have such information.” **(caregiver, a hospital attendant)***

The management of complications such as hydrocele in a patient gave an insight into how these factors can affect their decision to avail healthcare services related to lymphadenitis.

> *“He never used to go for consultation. I forcefully took him, and they did an ultrasound. He was diagnosed with ripe filaria. Then only we went for surgery, it took around forty to fifty thousand rupees. Then he took 6 months rest. Then only (the treatment) worked.” **(caregiver of vegetable seller in her 30s)***

#### Hopelessness from not getting cured

Because of all these barriers affecting the patients’ suffering, they experienced strong feelings of dissatisfaction about never getting cured of filariasis and ADLA. Different modes of therapy had failed in the end to improve the quality of life for some of the patients.

> *“There was a Homeopathic doctor in the colony. Many people used to say that the treatment is effective, but it didn’t work for me. There was another doctor who used to give some zaributi (plant-based) medicine, he told not take food throughout the day after taking the medicine, and to take curd in the evening. The whole day I had to starve. But nothing happened. Then I became tired of it and left it… the disease was not cured, so how one could be satisfied?” **(patient, Anganwadi worker for 15 years)***

This could adversely impact their mental well-being, affecting even the caregivers, who had spent a great proportion of their time caring for the disabled individuals. They had witnessed the pain and suffering of their loved ones, which rendered them disgruntled and hopeless about the whole situation.

> *“See, some disease has no cure, like AIDS. (It’s possible) only if you take precautions. I know this disease (Filariasis) can’t be cured. But treatment must cure the pain. Like, pain occurs as if it may kill you. So, to avoid that, government should do something, to reduce the pain.” **(caregiver, a hospital attendant)***

Hopelessness led to expectations for better intervention from the government. This situation points to the existing gap between guidelines and the reach of successful ground-level interventions of the MDA-MMDP programme, which had not managed to prevent the occurrence of filariasis in the community.

#### Not maintaining personal hygiene

Some patients in the community were not using soap to clean the affected limbs, even though a water supply was present in their household. Instead, they were using ashes to care for the infected leg. Another necessity was taking adequate rest. But most of them were either involved in doing work without any break or not fulfilling the basic requirements of foot care, skin care, exercise, and leg elevation.

> *“It becomes very difficult to do farming in the cold… I go without (slippers).” **(patient, farmer in his 40s)***

> *“This thing, I always tell her. That whenever you sit, wherever you sit, always have your legs raised up… She never listens.” **(caregiver, a hospital attendant)***

Maintaining hygiene is the most important need for ADLA patients who are suffering for prolonged periods of time. This aspect had been very much neglected by most of the respondents, which continued to add to the healthcare costs and burden of filariasis in the community.

#### Hampered activities of daily living

Patients reported difficulty in maintaining balance, walking, standing up or sitting down, or doing the basic household chores like cooking, washing, or cleaning. The main culprits were pain and swelling of the lower limbs, which hampered the normal movements of the patients. One of the patients expressed her plight in facing severe difficulty in doing chores at home, with no one to help her at all:

> *“I am not able to do cold related work. What else! …Like, work related with water, have to do it irrespective of other family members at home or not. Even if I worry, what can I do! If there are no daughters or daughter-in-law at home, one has to do it. At least cooking and cleaning have to be done, who will do all these things? No one is here.” **(patient, homemaker in her 70s)***

#### Psychosocial difficulty

Few respondents reported that they had decreased socializing in their neighbourhood because of physical difficulty. One respondent informed about financial difficulty with filariasis in the past while his children were still dependent. Though there was no stigma in the study community regarding filariasis and ADLA, some patients seemed to go into recluse or even delve into addiction once they were known to be afflicted with this disease.

A caregiver reported that the patient did not consume MDA given by ASHA and only took medicine from a private facility, even though his whole family took MDA. He also didn’t care for his lymphedema in the lower limb. However, since he had filariasis, the main problem reported was his addiction to alcohol:

> *“He stays outside, goes to village. Now he goes astray, and then drinks (alcohol). Now when he drinks, only that can bring some relief. This is the main problem, nothing else. If only his alcohol addiction was managed, then there would be no problem.” **(caregiver of vegetable seller in her 30s)***

Perhaps the consumption of alcohol by this patient reflected a much deeper problem in this setting. The feeling of never getting out of the ‘abyss’ of filariasis can negatively impact the quality of life of whole families, not just the patients.

### Healthcare system related factors (theme 3)

#### Lack of capacity building for grassroots workers

The reason healthcare workers are not taking adequate care of patients could be attributed to infrequent training regarding the disease. One grassroots healthcare worker, when asked if she had attended any meeting or training on filariasis, replied that she had attended only one meeting regarding managing filariasis in the community:

> *“Yes, long time ago, but I don’t remember much about it now… I tell them (patients) to keep wet warm cloth over the limb, but it has not been told how long they should do it.” **(health care worker)***

Unfortunately, there had been no formal training programme for capacity building of the healthcare workers on the management of filariasis and ADLA in the community. The only time ‘meetings’ were held were on “surveys regarding how many patients are positive” for filariasis.

#### Lack of receipt of regular incentives for grassroots workers

One of the major problems in managing patients with filariasis was a lack of motivation and disappointment, which stemmed from the fact that healthcare workers were not getting any incentives for taking filariasis patients to health facilities.

> *“Yes, if we will get something for filariasis obviously we would pay more attention to those patients and will bring them to BPHC.” **(health care worker)***

> *“No incentive for taking patient to hospital. We get incentive during program, for house to house, mass drug administration in people starting from more than two year old.” **(health care worker)***

On the contrary, the ***block health manager*** of the same block told a different story:

> *“Yes, a hundred rupees for bringing patient… yesterday all ASHA who brought patient, they all got hundred rupees. They get incentives for all work.”*

Because of the lack of regular incentives for healthcare workers, some patients reported very few visits from the grassroots workers. Even if they mobilized the patients, all they did was give the MMDP kits to the patients at the BPHC every Monday. They did not have any information on how to clean the ADLA-affected limbs to decrease the chances of ADLA attacks.

#### Unavailability of laboratory diagnosis of filariasis at health facility

Laboratory diagnosis of LF by examination of blood at night is an important follow-up method for detecting any active traces of infection in patients. The investigation by microscopic examination was not available at any nearby health center or even the BPHC, while management of the disease was based mainly on clinical diagnosis. The need for laboratory diagnosis and advice for management of the disease sequelae was very much felt at the village level by the patients and their relatives.

> *“No such test was there. As you people are saying, there is no treatment for filaria. Doctor said, there’s no any internal problem. So, we didn’t do anything else. They didn’t tell anything as such (about taking care of swollen leg).” **(caregiver, a hospital attendant)***

#### Unavailability of management of complications of filariasis at health facility

Treatment of complications was not available at the BPHC. On OPD visits, they were not given advice regarding how to take care of the affected limb at home. Thus, patients had to travel long distances to a tertiary health center to avail of such services, which made even the healthcare workers apprehensive.

> *“Most people don’t go to block, even at my home there are two patients of filariasis but they don’t go to hospital. It would be better if more facilities for treatment will be provided at the center!” **(health care worker)***

This was a dangerous precedent for the growing burden of complications in the community. An urgent need was felt to scale up the services and initiate minor surgical and dressing procedures in the peripheral health facilities of the area for care of the affected limbs and complications like ADLA and hydrocele.

#### Inconsistent drug supply in the community and health facility

The treatment component of filariasis was mostly symptomatic among the respondents. diethylcarbamazine, ivermectin, and albendazole had not been administered to many patients because of their inconsistent availability at the BPHC. A relative remembered that besides not getting tested in a laboratory, the patient also did not receive the main chemotherapeutic management of the disease from a tertiary hospital, which was quite alarming.

> *“One year back, I sought treatment (for patient). We got only calcium. Nothing else. Calcium and BP.” (caregiver, a hospital attendant)*

The ***block health manager*** though, recounted differently:

> *“Filariasis is very common here. Yesterday we distributed kits, as many patients came. Patient presents with fever, cough, swelling of limbs. Even if slight swelling is there, we start DEC 6mg/kg. Suppose 70kg male presents with swelling then dose will be 420mg. So we give 100mg thrice daily with Levocetirizine which prevents drug allergy.”*

The healthcare workers took the patients to the BPHC, where they received adequate treatment only when there was a supply of diethylcarbamazine and albendazole. There could be certain areas in the community that had been missed during the MDA implementation too. One patient in the study area complained about not receiving the MDA drugs from the healthcare workers during the biannual deworming days.

#### No financial assistance

Most of the patients had to buy preventive measures like long-lasting insecticide nets (LLINs) from shops, although they were supposed to receive them from the public distribution system or from the health facility under NVBDCP. But both the healthcare workers and the block health manager corroborated that the BPHC did not receive any supply for the LLINs.

> *“Mosquito nets they (patients) have to purchase.” **(health care worker)***

> *“They do not get LLINs from here. They get from government supply. They just get medicines, bath tub, mug, antifungal, soap and povidone iodine from here.” **(block health manager)***

This was an added burden for those who could not even afford three meals every day, as the majority of the filariasis patients in the study area belonged to very poor socioeconomic class. Both patients and grassroots workers complained that the afflicted are not benefited by any means by attending the BPHC. Because of the dearth of so many services in government health facilities, some patients were forced to resort to treatment in the private sector.

> *“We went (to doctor) in 2016 only. Did ultrasound, and then did operation (for hydrocele). Yes it’s private. We take medicines from private. They give medicine of hundred or fifty rupees, only then he becomes fine, otherwise not.” **(caregiver of vegetable seller in her 30s)***

All these factors contributed to increased out-of-pocket expenditures. Households with patients with LF can incur huge costs, especially if they are in the low-income group or if they are seeking medical care in the private sector instead of a government health facility.

## Discussion

LF is a debilitating disease that is most responsible for permanent disfigurement in the afflicted, making it the second-most common cause of long-term disability worldwide [6]. Patients repeatedly suffer in agony due to symptoms that vary with temperature and humidity [7]. A study in Ghana showed that the majority of the symptoms occurred in the rainy season, as the maximum infection of filariasis happens in the period of minimum vector density due to flushing of the breeding places, whereas the attacks are less prevalent in the dry season [8]. Similarly, in the North Indian and East Indian states, several factors increase the probability of filarial transmission in the summer. Tropical climate, poorly maintained drainage, and increased vector-human contact attributed to using fewer clothes and sleeping in open or outside the room at night are responsible [9]. Only two of our respondents suffered in the winter, compared to five in the summer. But a study done in Tamil Nadu, South India, has shown no significant variation between the summer and winter seasons [10]. The survival of the vector *Culex quinquefasciatus* was less in the summer, and infective stage larvae were not encountered until the early monsoon. The parous mosquitoes have been recorded highest in the winter months (November-March) in this part of India, leading to maximum transmission that declined with a rise in temperature [11]. Geographical differences seem to be playing an important role; however, our respondents showed mixed variation. The best way to prevent LF is to avoid mosquito bites, followed by the biannual MDA program, while the WaSH strategy has been increasingly recognized as crucially important in the prevention of morbidity and disability for filariasis-related lymphedema [5, 12].

In our study, the respondents had low awareness regarding treatment and prevention of the disease, resulting in low MDA adherence. In an endemic town in Nigeria, 85% did not know about MDA, and 36% did not know its role in preventing LF [13]. Joseph et al. conducted a study among health workers in Karnataka, India, where they found poor awareness scores in 37.2%, average in 57.7%, and good scores in only 5.1% of participants [14]. The patients might fear its side effects, remain absent during the time of distribution, not consume the drug as they forget or misplace it, or suffer from a concurrent illness [15]. They also feel mounting hopelessness that lymphedema is incurable, so there is no use in diligently following management practices once they contract filariasis. However, increased community participation in WaSH needs to gain traction. Patricia Maritim et al. have concluded that adherence and health-seeking behaviour were even driven by negative beliefs and myths about the disease and a lack of awareness about available homecare morbidity kits and WaSH facilities [16]. WaSH programmes can also help reduce the breeding grounds for the mosquito vectors that breed in the open sewers [17]. In a study by Mues et al. in Odisha in 2014, overall compliance with lymphedema management techniques due to a community-based management programme was negatively associated with difficulty accessing water and soap, emphasizing that lymphedema management is incomplete without WaSH facilities [18]. In Nepal, even though patients suffering from lymphedema had home-based care as their first treatment, their first point of contact were the traditional healers, only later seeking Ayurvedic or modern hospital-based treatments during complications [19]. Even unfavourable interactions between the provider and the client reduce acceptability. The low health-seeking behaviour among the patients for modern medicine found in this study has been attributed to low trust in government facilities, leading to failure in cure and mounting hopelessness. Sometimes, misconceptions and rumours can spread like wildfire, obstructing the programme and community participation [20].

The other problems in the study were that patients and caregivers were unable to work productively, leading to loss of earnings from reduced work hours, catastrophic and inappropriate health costs leading to debt, not being able to pay for the schooling of children in the family, and an overall reduced societal role. The younger the cohort, the greater the economic losses that accumulate over their years of productivity [17]. This is because patients whose lymph vessels are already damaged experience lymphedema and ADLA episodes for the rest of their lives. They also undergo increased travel costs, coupled with lost productivity in both paid and unpaid labour [21]. All these surely affect their mental health as well, as evidenced by filariasis being related to depression and other psychosocial difficulties [22]. The social stigma of lymphedema can cause barriers both to adherence and health-seeking behaviour, as seen in several studies [5], though this study did not find any such complaints.

The filariasis elimination programme had faced severe implementation challenges for years. The innovative efficiency was initially slow, and the disease impact on endemic regions was rarely focused on. Thus, the WHO expanded its technical support to address these issues from 2017 until 2019. Microplanning and social mobilization might have improved in the case of MDA dispensing, but training about home-based care was still lacking. The two healthcare workers in our study had attended only refresher meetings held on annual surveys or before the distribution of MDA. There was no formal training on how to manage lymphedema. It seems the state health authorities have put more importance on the transmission of microfilariae but side-lined how to improve the quality of life in patients with severely affected lymphatics and cutaneous inflammation. Once established, the disease is irreversible even after treatment or spontaneous death of the microfilariae, which promotes insidious progression [23]. Initial ADLA attacks due to bacterial proliferation worsen the lymphedema, leading to elephantiasis. This creates a vicious cycle favouring more such attacks due to a lack of local hygiene [24]. Implementation at the administrative level is only strengthening the MDA administration and not the second pillar of the elimination strategy, which is managing the morbidity [25].

Many studies have elaborated on the physical, social, psychological, sexual, and economic problems not only caused by the deformities but also by the acute febrile episodes [26–28]. Even then, the administration had to face several problems. During the rollout of IDA round-1 in Bihar in 2019–2020, there was a local strike by ASHAs at booths established in Anganwadi centres and schools. Only 58–61% of the total population had consumed the triple drug regimen, which was soon interrupted when Anganwadi workers also joined the strike [29]. During the study period, there were subsequent strikes occurring at the BPHC due to the non-payment of any incentives by the government over a prolonged period. This demotivated the grassroots healthcare workers, who were already burdened with mobilization and drug distribution under different community health programs. Hydrocelectomy surgery is not conducted at the BPHC, which forced one patient to a private clinic, resulting in catastrophic expenditure. There, only MMDP kits were distributed every Monday, with no training on how to use them to prevent ADLA. Lessons can be learned from Kerala State regarding this. The State Health Administration of Kerala conducted intensive training sessions for doctors and staff nurses from each Taluk hospital to provide care for lymphedema patients. They were given training in MMDP, including information, hands-on demonstrations, and specific lymphedema care plans. This increased the registration of previously unidentified lymphedema patients. Undertaking a policy decision to achieve elimination with MMDP in addition to MDA is the key component of that strategy, which should be the goal for endemic districts in all of India [3].

## Conclusions

The main barriers that affected the quality of life of lymphedema in LF patients were the inherent nature of the disease, including symptoms and seasonality, patient related factors, and factors due to healthcare system delivery. These grassroots problems are essential for understanding the impact that lymphedema and ADLA have on patients’ quality of life and highlighting what to really address to increase home-based care for LF. Accessibility to WaSH, regular training of home-based care, increasing the capacity and motivation of grassroots workers, and the generation of in-depth awareness among the patients are required to achieve the elimination of filariasis, with MMDP as a key component of that strategy for endemic districts across the whole country.

## Supporting information

Additional file 1: Text S1. Interview guide

Additional file 2: Table S1. Content analysis of the categories in the study

Additional file 3: Annexure S1. Participant information sheet. Annexure S2. Participant informed consent form

## Data Availability

All the quotes, codes, and categories described in the manuscript will be available to any scientist wishing to use them for non-commercial purposes. The confidentiality of the participants will be maintained.

## Abbreviations

LF: lymphatic filariasis
MMDP: morbidity management and disability prevention
MDA: mass drug administration
WaSH: water, sanitation and hygiene
WHO: World Health Organization
ADLA: acute dermato-lymphangio-adenitis
ASHA: accredited social health activist
BPHC: block primary health center
ANM: auxiliary nurse midwife
NVBDCP: National Vector Borne Disease Control Programme
LLIN: long-lasting insecticidal net

## Acknowledgements

The authors thank the Department of Community and Family Medicine, AIIMS Patna, especially Mr. Nitin Kumar Singh, Medical Social Service Officer, for field coordination during the study.

## Authors’ contributions

PK, SA and DB contributed equally to this work in framing the concept, background and protocol of the study, data collection and manuscript drafting. RR contributed to data collection, analysis, manuscript drafting, and reviewing. DB and BS contributed equally to this work in data analysis and manuscript reviewing. All authors read and approved the final manuscript.

## Ethics approval and consent to participate

Ethical approval was obtained from the Institutional Ethical Committee of the All India Institute of Medical Sciences, Patna (AIIMS/Pat/IEC/2019/351). Informed consent was obtained from all participants for their interview to be recorded before beginning the conversation (Additional file 3: Annexure S1 and Annexure S2). The participants were first explained in detail about the nature of the study, following which it was conveyed that participation is voluntary and they are free to withdraw from the study at any time, without citing any reason. There were also no risks involved in the study, which the participants were informed about. The consent forms, audiotapes, and transcripts were stored on a password-protected computer, which was only accessible to the researchers. Any report, presentation, or publication resulting from this research does not contain any information, such as names, that could identify the participants or their relatives.

## Consent for publication

Not applicable

## Supplementary Information

**Additional file 1: Text S1.** Interview guide

**Additional file 2: Table S1.** Content analysis of the categories in the study

**Additional file 3: Annexure S1.** Participant information sheet.

**Annexure S2.** Participant informed consent form

