## Additional file 1: Text S1. Interview guide for "*“Whenever I tell her to wear slippers, she turns a deaf ear. She never listens”* : a qualitative descriptive research on the barriers to basic lymphedema management and quality of life in lymphatic filariasis patients in a rural block of eastern India"

1. Tell me about yourself:
   a. **Prompts:** where are you from? How long have you lived there? How many people in your family? What kind of work do you do?
2. How has this past year been for you?
3. Tell me when you first found out about your lymphatic filariasis.
   a. **Prompts:** how many years back did you find out? how did you find out? How did you cope? How did you inform family members or community members, if at all? How has the diagnosis affected you, if at all?
   b. When did the lymphedema started? How did you cope?
4. Have you been treated for filariasis?
   a. **Prompts:** How were you been treated? Where did you go for treatment? What advices have been given there?
5. Do you know how you can prevent the progression of disease?
   a. **Prompts:** Have you experienced episodes of fever and pain in affected limb in past year? How frequently it was used to happen in the past year? What can you do to halt disease progression?
6. What factors do you think contribute to a good quality of life?
   a. **Prompts:** Why are these factors important? Well-being? Appreciation of life? Family life? Health? Living environment?
7. How do you feel about your quality of life?
   a. **Prompts:** How is your living situation? How has the swelling affected your personal relationship, social relationship and job?
8. How do you feel about your care and treatment?
   a. **Prompts:** Can you take care of yourself? How family members helped you in daily life? What, if anything, would you change anything about your care and treatment?
9. What is preventing you to take care for swelling?
   a. Do you have soap and access to water in home? Can you wash your limbs your own? Do other family members help you to wash? Did you do exercise? Did you elevate your limb?
