## Additional file 2: Table S1. Content analysis of the categories in the study for "*“Whenever I tell her to wear slippers, she turns a deaf ear. She never listens”* : a qualitative descriptive research on the barriers to basic lymphedema management and quality of life in lymphatic filariasis patients in a rural block of eastern India"

| Categories | n (%) of total codes |
| --- | --- |
| Symptoms and signs of the disease | 55 (35.3) |
| Seasonal factors | 14 (9.0) |
| Low awareness in patients and relatives | 14 (9.0) |
| Low adherence | 12 (7.7) |
| Low health-seeking behaviour in modern medicine | 5 (3.2) |
| Hopelessness from not getting cured | 3 (1.9) |
| Not maintaining personal hygiene | 18 (11.5) |
| Hampered activities of daily living | 5 (3.2) |
| Psychosocial difficulty | 5 (3.2) |
| Lack of capacity building for grassroots workers | 3 (1.9) |
| Lack of receipt of regular incentives for grassroots workers | 4 (2.6) |
| Unavailability of laboratory diagnosis of filariasis at health facility | 6 (3.8) |
| Unavailability of management of complications of filariasis at health facility | 2 (1.3) |
| Inconsistent drug supply in the community and health facility | 4 (2.6) |
| No financial assistance | 6 (3.8) |
| TOTAL | 156 (100) |
