## Additional file 3: Annexure S1. Participant information sheet. Annexure S2. Participant informed consent form for "*“Whenever I tell her to wear slippers, she turns a deaf ear. She never listens”* : a qualitative descriptive research on the barriers to basic lymphedema management and quality of life in lymphatic filariasis patients in a rural block of eastern India"

**Title: Assessment of Quality of life and Barriers to Basic Lymphedema Management in Lymphatic Filariasis patients: A Mixed-method study from rural Patna, Bihar**

The main objectives of the study are to assess perceived quality of life and to identify the barriers of utilizing the basic lymphedema management in filarial lymphedema patients. You will be asked to rate your quality of life in physical, social and psychosocial domains. Further, an in-depth interview exploring the barriers of lymphedema care and management will be inquired. The in-depth interview will be audio-taped. The study will contact you for only one time. You can choose whether to participate in the focus group and stop at any point of time. Although the group discussion will be recorded your responses will remain anonymous and no names will be mentioned in the final report. The duration of discussion will be of 45-60 minutes in which your participation is purely voluntary.

**Risks or disadvantages to participants:** There are no other risks involved that might affect you directly.

**Any benefits to participants:** There are no direct benefits, but by taking part, you will help us

In understanding perceptions of quality of life and problems faced by you due to

lymphedema that potentially help in improving care management in the future.

**Data management:** Any information that can identify you personally such as your name or

address shall be removed. After the research has been completed, the anonymized data

collected during the study period shall be stored as per local regulations.

**Annexure S2:	Participant informed consent form**

Title: Assessment of Quality of life and Barriers to Basic Lymphedema Management in Lymphatic Filariasis patients: A Mixed-method study in rural Patna, Bihar.

Research Institute: All India Institute of Medical Sciences, Patna ……...Tel No: 6122-452106

The contents of the information sheet that was provided have been read carefully by me/ explained in detail to me, in a language that I comprehend, and I have fully understood the contents. I confirm that I have had the opportunity to ask questions.

The nature and purpose of the study and its potential risks / benefits and expected duration of the study, and other relevant details of the study have been explained to me in detail. I understand that my participation is voluntary and that I am free to withdraw from the study at any time, without giving any reason, without my medical care or legal right being affected. I understand that the interview will be audio-taped for future report preparation.

I agree to take part in the above study.

--------------------------------------------	Date:
(Signatures / Left Thumb Impression)	Place:

Name of Participant: _________________________Son/Daughter/spouse of: _________________
Complete postal address:___________________________________________________________

This is to certify that the above consent has been obtained in my presence.

------------------------------	Date:
Signatures of the Principal Investigator	Place:

**1) Witness – 1	2) Witness – 2**

**……………………………………….. …….……………………………….**

Signature	Signature

Name	Name

Address	Address
